# Infants and young children generate more durable antibody responses to SARS-CoV-2 infection than adults

**DOI:** 10.1101/2023.04.10.23288360

**Authors:** Devyani Joshi, Lindsay E. Nyhoff, Veronika I. Zarnitsyna, Alberto Moreno, Kelly Manning, Susanne Linderman, Allison R Burrell, Kathy Stephens, Carson Norwood, Grace Mantus, Rafi Ahmed, Evan J. Anderson, Mary A. Staat, Mehul S. Suthar, Jens Wrammert

## Abstract

Since the emergence of SARS-CoV-2, research has shown that adult patients mount broad and durable immune responses to infection. However, response to infection remains poorly studied in infants/young children. In this study, we evaluated humoral responses to SARS-CoV-2 in 23 infants/young children before and after infection. We found that antibody responses to SARS-CoV-2 spike antigens peaked approximately 30 days after infection and were maintained up to 500 days with little apparent decay. While the magnitude of humoral responses was similar to an adult cohort recovered from mild/moderate COVID-19, both binding and neutralization titers to WT SARS-CoV-2 were more durable in infants/young children, with Spike and RBD IgG antibody half-life nearly 4X as long as in adults. The functional breadth of adult and infant/young children SARS-CoV-2 responses were comparable, with similar reactivity against panel of recent and previously circulating viral variants. Notably, IgG subtype analysis revealed that while IgG1 formed the majority of both adults’ and infants/young children’s response, IgG3 was more common in adults and IgG2 in infants/young children. These findings raise important questions regarding differential regulation of humoral immunity in infants/young children and adults and could have broad implications for the timing of vaccination and booster strategies in this age group.

## Introduction

More than three years since the SARS-CoV-2 virus was first detected in humans, it continues to cause severe morbidity and mortality worldwide (1, 2). Durable immune responses against SARS-CoV-2 are crucial for prevention of severe disease and for protection against the continuously emerging viral variants (3). The breadth and durability of the immune response has been extensively studied following infection, as well as after primary and booster series of vaccination, in both adults and older children (4–10). However, the immune response following SARS-CoV-2 infection in infants and very young children remains poorly understood. Although approximately one fourth of infants infected with SARS-CoV-2 are asymptomatic and there are few reported deaths, severe COVID-19 is more common in young infants as compared to older children (11–13). The immune system of infants starts to develop in the first few months after delivery (14). Very little is known about how the immature immune system of an infant or young child reacts to SARS-CoV-2 infection, and how the infection in those children in turn impacts the development of the immune system. Even less is known about the durability of immunity, given the difficulty of designing and executing multi-sample longitudinal studies in these age groups. Thus, evaluation of the initial magnitude and the long-term durability of infection-induced immune responses in infants/young children is critical. Moreover, a deeper understanding of the breadth of humoral immune responses against continuously emerging viral variants in infants/young children is vital for optimizing the timing of current vaccination strategies in this age group (15, 16).

In this study, we present findings from the IMPRINT study; a prospective, longitudinal birth cohort of influenza and SARS-CoV-2 infection and vaccination in early life conducted at Cincinnati Children’s Hospital Medical Center. IMPRINT participants provide weekly parent-administered mid-turbinate nasal swabs, allowing for real-time identification of SARS-CoV-2 infected infants and young children. Multiple blood samples were obtained from these infants and children over a period of up to 500 days after their initial SARS-CoV-2 positive swab. This pediatric cohort was then compared to an adult cohort on which we have previously reported, which collected blood samples from patients with PCR confirmed SARS-CoV-2 infection that were followed for up to 350 days post infection (5). Herein we report that while the initial magnitude of the SARS-CoV-2 specific antibody response in adults and infants/young children was comparable, the titers in our infant cohort were maintained over the study period, while in adults the titers declined with a half-life of 180 days (as previously reported by us and others) (5, 17, 18). Despite this major difference in terms of durability of humoral immunity, the functional breadth of these responses was similar in adults and infants/young children, since the breadth of reactivity against a panel of viral variants was essentially identical between these groups. Our findings show significant differences in the maintenance of antibody responses in adults and infants/young children and suggest that booster vaccination scheduling strategies in infants/young children may need further evaluation.

## Results

### Study cohort

We included a total of 23 infants/young children and 19 adults in the study cohort. The infants/young children, recruited at Cincinnati Children’s Hospital, were followed since birth and up to 500 days after their first positive PCR result for SARS-CoV-2. The COVID-19 confirmed adult patients were previously enrolled in our longitudinal study of COVID-19 immune durability (5), and followed for up to 350 days post-infection. Of the adults, a majority (84 %) were non-hospitalized, and none had severe symptoms. Infants/young children were infected during April 2020 – January 2022, and adults were infected during March – April 2020 (Supplementary Figure 1) (19, 20). The demographics and baseline characteristics of these cohorts are described in **Table 1**. The infant cohort was 57 % females and 43 % males and between 40 and 1441 days old (median age, 249 days). The adult cohort was 53 % females and 47 % males and between 20 and 77 years (median, 54 years).

**Table 1:** Cohort demographics

|  |  | Infants/young children | Adults |
| --- | --- | --- | --- |
| Sex | Females (%) | 13 (57%) | 10 (53%) |
|  | Males (%) | 10 (43%) | 9 (47%) |
| Median age (range) |  | 249 d (40 – 1441 d) | 54 y (20 – 77 y) |
| Samples collected |  | February 2020 – April 2022 | May 2020 – March 2021 |
| COVID-19 infection |  | April 2020 – January 2022 | March 2020 – April 2020 |
| Patients requiring hospitalization |  | None (0 %) | 3 (16 %) |
| Severe symptoms |  | None (0 %) | None (0 %) |

### Durable antibody responses in infants/young children after SARS-CoV-2 infection

The magnitude and durability of antibody responses after SARS-CoV-2 infection have not been carefully studied in infants/young children. Here we evaluated the longitudinal plasma antibody responses against several SARS-CoV-2 antigens (Spike, RBD and NTD) pre-infection, during acute infection, and following recovery in 23 SARS-CoV-2 infected infants and young children using the Meso Scale Discovery electrochemiluminescence multiplex assay (MSD). All infants/young children exhibited measurable IgG and IgA responses against Spike, RBD, and NTD after infection, while IgM responses were negligible not only prior to infection, but also at acute infection and convalescence stages in most individuals (data not shown). Pre-infection levels of IgG and IgA against SARS-CoV-2 spike (**Figure 1A and D**), spike receptor binding domain (RBD) (**Figure 1B and E**), and spike N-terminal domain (NTD) (**Figure 1C and F**) were below the limit of detection, excluding an impact by maternal antibodies. The SARS-CoV-2-specific IgG and IgA levels increased rapidly during acute infection, with subjects generating detectable antibody responses as early as day four and peaking approximately 30 days post-infection. The antibody titers were maintained following recovery for up to 500 days after the positive PCR test with little apparent decay.

**Figure 1:**
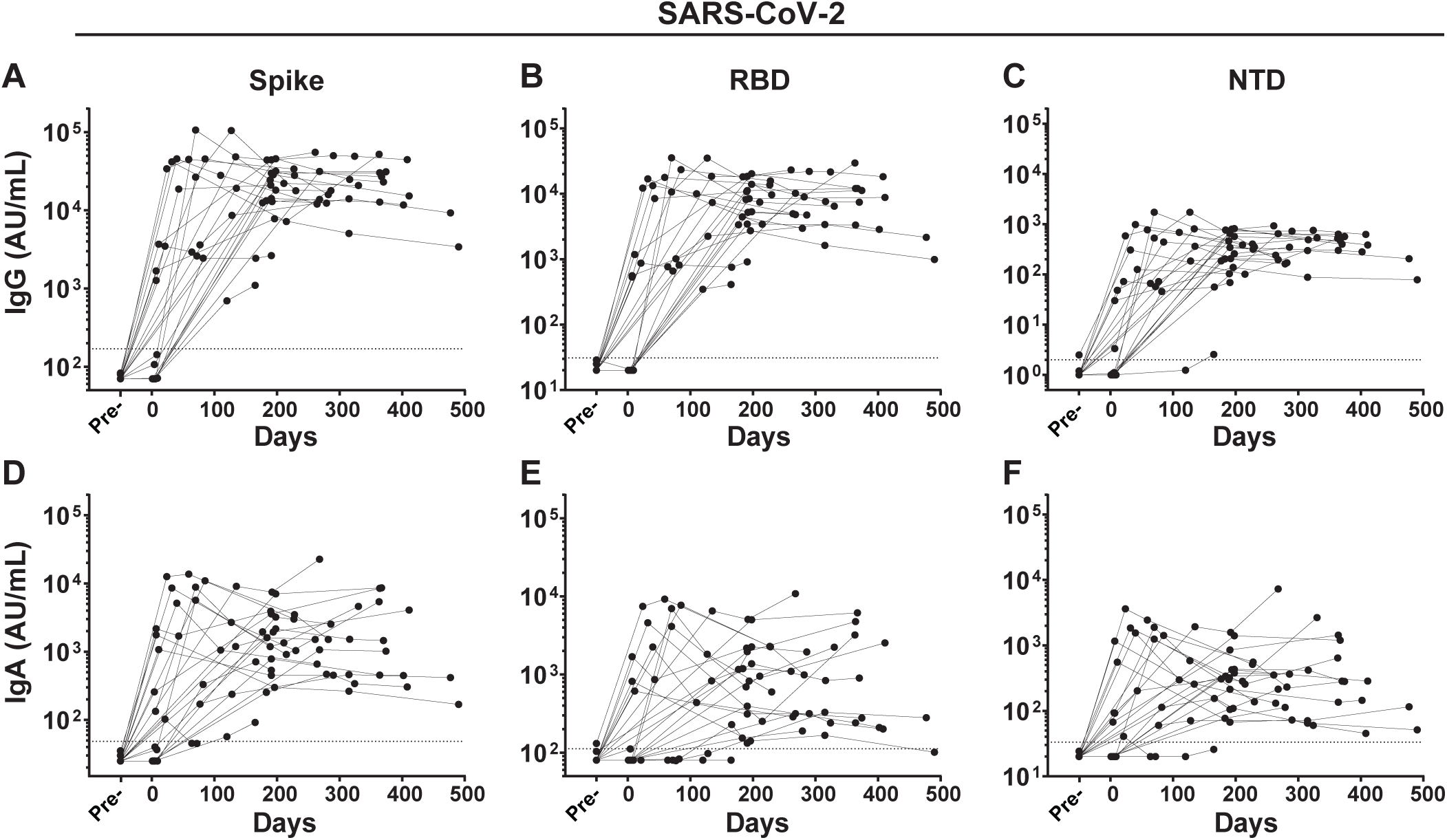
Infants/young children generate durable antibody responses to SARS-CoV-2 antigens following infection. IgG (A – C), and IgA (D – F) antibodies reactive to SARS-CoV-2 spike (A, D); spike receptor binding domain (RBD) (B, E); and the spike N-terminal domain (NTD) (C, F) were measured in duplicate by an electrochemiluminescent multiplex immunoassay and reported as arbitrary units per ml (AU/mL) as normalized by a standard curve. Longitudinal antibody titers of SARS-Co-2 infected infants/young children (n = 23) are plotted over days post positive PCR result. The dotted line represents the limit of detection, defined as the mean value pre-infection + 3SD.

### SARS-CoV-2 infection results in increased antibody titers against the epidemic coronaviruses MERS-CoV and SARS-CoV-1 in infants/young children

We further evaluated the IgG antibody titers against the epidemic coronaviruses SARS-CoV-1 and MERS-CoV spike protein. The IgG binding antibody titers against both SARS-CoV-1 and MERS-CoV spike protein increased during the acute phase of SARS-CoV-2 infection. IgG titers peaked approximately 30 days post-infection and were maintained with little to no apparent decay for up to 500 days post infection. (**Figure 2A**). Given that none of the infants/young children could have been exposed to these coronaviruses, the increase in the IgG antibodies most likely represents cross-reactive antibodies directed to SARS-CoV-2 spike epitopes which are largely conserved between SARS-CoV-1, MERS-CoV and SARS-CoV-2 (21). This shows a heightened immunity to the epidemic coronaviruses upon SARS-CoV-2 infection in infants/young children.

**Figure 2:**
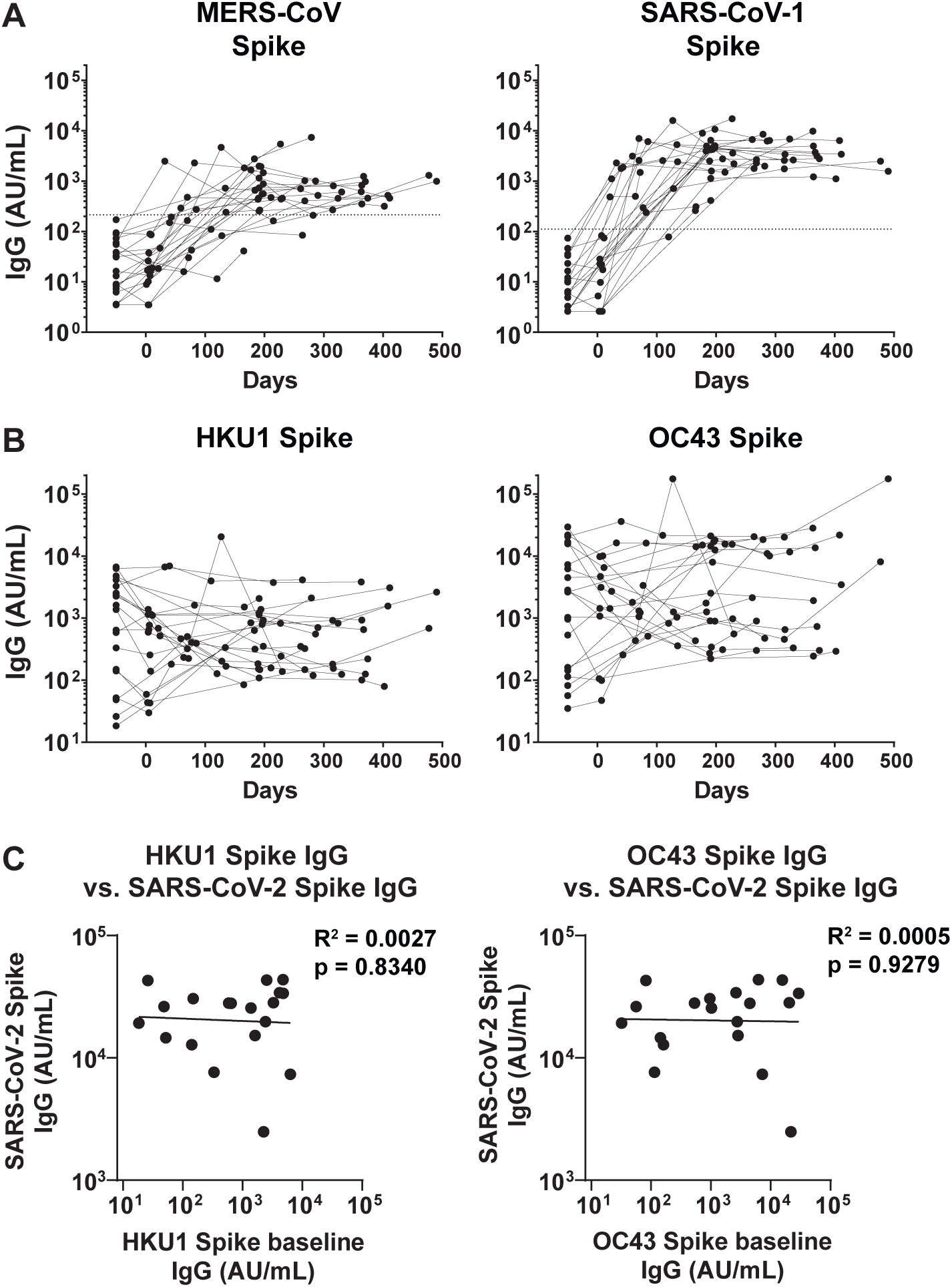
**Longitudinal antibody responses against other coronavirus spike proteins in infants and their correlation to SARS-CoV-2 spike binding antibody response**. IgG antibodies reactive to the (A) previous epidemic coronaviruses, MERS-CoV and SARS-CoV-1 spike protein, and (B) common human beta-coronaviruses, HKU1 and OC43 spike protein were measured in duplicate by an electrochemiluminescent multiplex immunoassay and reported as arbitrary units per ml (AU/mL) as normalized by a standard curve. Longitudinal antibody titers of SARS-CoV-2 infected infants/young children (n = 23) are plotted over days since the positive PCR result. (C) Pre-SARS-CoV-2 infection IgG binding antibody titers against the spike proteins of currently circulating human beta coronaviruses, HKU1 and OC43, showed no correlation with the convalescent titers against SARS-CoV-2 spike in infants/young children. Coefficient of determination (R^2^), and significance were determined by linear regression analysis.

### Pre-existing IgG antibody titers against the seasonal β-coronaviruses HKU1 and OC43 spike protein do not impact antibody responses after SARS-CoV-2 infection

It has been postulated that pre-existing antibody responses against seasonal beta-coronaviruses might impact the responses against SARS-CoV-2. Since we had access to pre-infection samples for this cohort, we next evaluated the IgG binding antibody response to the common human coronaviruses HKU1 and OC43 in the pre-infection, acute infection, and convalescent plasma samples from 23 SARS-CoV-2 infected infants/young children using the MSD assay. Overall, IgG titers against the spike proteins of HKU1 and OC43 were largely unaffected by the SARS-CoV-2 infection (**Figure 2B**). More importantly, post-infection titers against the SARS-CoV-2 spike protein showed no correlation to the baseline (pre-infection) titers against the HKU1 and OC43 spike protein (**Figure 2C**) indicating that the level of pre-existing antibodies against common human coronaviruses did not impact the anti-SARS-CoV-2 antibody response following infection in infants and young children.

### Infants and young children show a more durable binding antibody response upon SARS-CoV-2 infection as compared to adults

An important aspect of serological immunity is the durability of protection induced by infection. To compare how the durability of antibody responses in infants/young children may differ from that in adults, we compared these results to a cohort of adult patients on which we have previously reported (5). The IgG responses to SARS-CoV-2 spike, RBD and NTD in infants and young children were found to show minimal to no decay over time, with an estimated half-life of 800 (95 % confidence interval [95 % CI] [222, ∞]), 775 (95 % CI [263, ∞]), and 828 (95 % CI [299, ∞]) days respectively, as estimated by the exponential decay model (**Figure 3A-C**). In contrast, IgG antibodies specific to SARS-CoV-2 spike, RBD and NTD in adults were found to decay much faster, with an estimated half-life of 187 (95 % CI [141, 281]), 178 (95 % CI [134, 264]), and 204 (95 % CI [149, 323]) days, respectively (**Figure 3E, 3F, and 3G**). We also calculated the antibody decay using the power law decay model, which assumes that decay rates decrease over time (5). The half-lives estimated by the power law model at day 150 for IgG antibodies binding to SARS-CoV-2 spike, RBD, and NTD in the infants/young children were 1964 (95 % CI [204, ∞]), 2986 (95 % CI [492, ∞]), and 14190 (95 % CI [192, ∞]) days, respectively. The longer half-lives as estimated by the power law model indicate that the decay rate of the IgG antibodies declines over time, and that the concentration of the antibodies starts to stabilize. In the adults, the half-lives of the IgG antibodies binding to spike, RBD and NTD, as estimated by the power law model at day 150, were 405 (95 % CI [250, 915]), 356 (95 % CI [228, 738]), and 450 (95 % CI [268, 1128]) days respectively.

**Figure 3:**
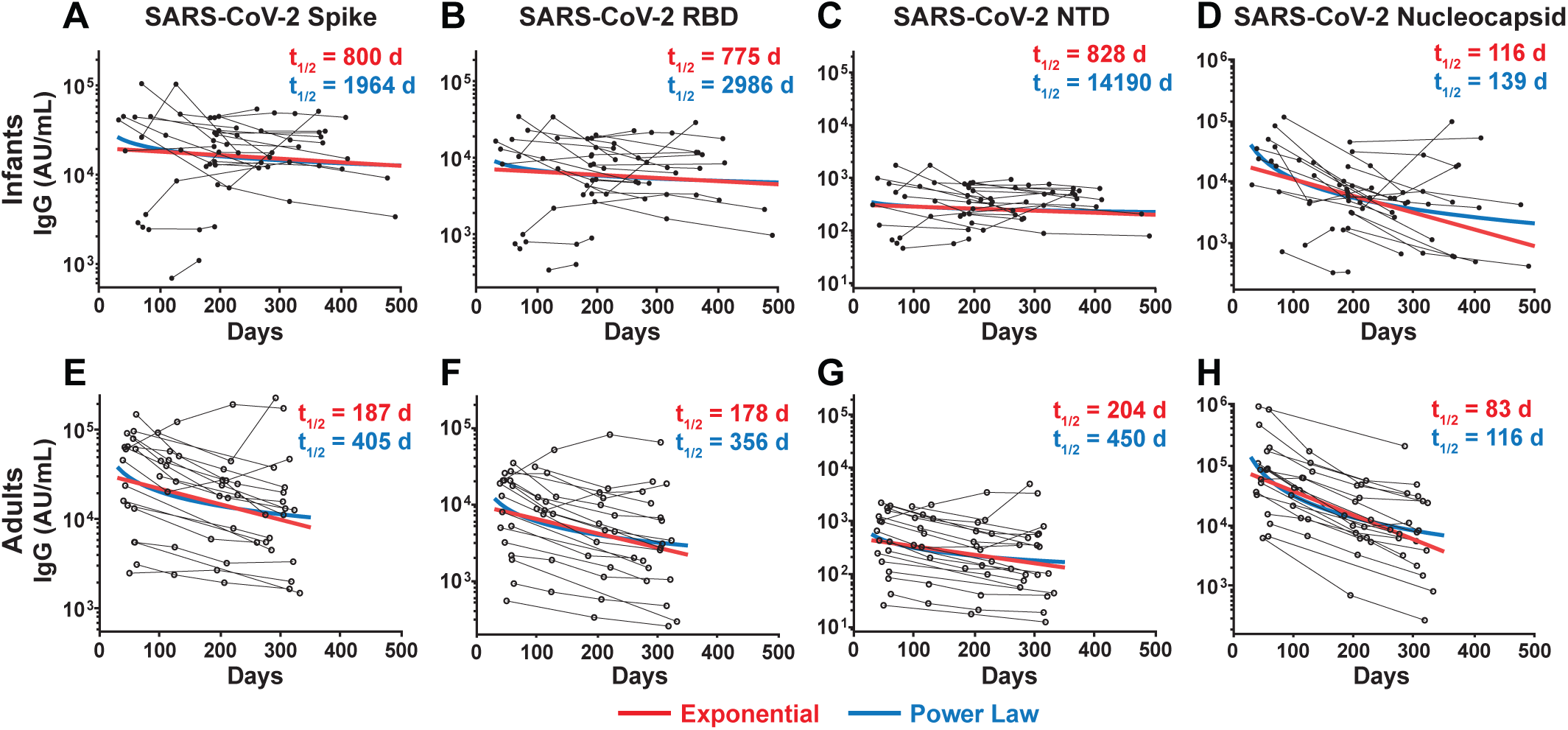
**Durability of SARS-CoV-2 spike- and nucleocapsid-binding antibody responses in infants/young children and adults.** IgG antibodies reactive to SARS-CoV-2 spike (A, E); spike RBD (B, F); spike NTD (C, G); and nucleocapsid (D, H) were measured in duplicate by an electrochemiluminescent multiplex immunoassay in infants/young children (A – D), and adult (E – H) donors and reported as arbitrary units per ml (AU/mL) as normalized by a standard curve. The half-lives and decay curves of the IgG antibodies estimated by the exponential model are shown in red; and the half-lives and decay curves of IgG antibodies estimated by the power law model (at day 150) are shown in blue.

As we have reported earlier in adults (5), anti-nucleocapsid IgG antibodies were found to decline more rapidly compared to antibodies to other SARS-CoV-2 antigens. Interestingly, we observed a similar fast decay in infants/young children, with the decay rate not statistically different between the two groups (p = 0.77 for the exponential decay model and p = 0.9 for the power law decay model, the Wald test). The half-lives of IgG antibodies binding to nucleocapsid protein in infants/young children as calculated by the exponential and power law models were 116 (95 % CI [80, 216]), and 139 (95 % CI [83, 388]) days respectively (**Figure 3D**). The half-lives of IgG antibodies binding to nucleocapsid protein in adults as calculated by the exponential and power law models were 83 (95 % CI [73, 96]), and 116 (95 % CI [97, 144) days respectively (**Figure 3H**). Taken together, these results suggest that infants and young children show a more durable binding antibody response to spike protein antigens and comparable durability of binding antibody response to nucleocapsid protein upon SARS-CoV-2 infection as compared to adults.

### Infants/young children and adults show differences in IgG subclass distribution after SARS-CoV-2 infection

To better understand the infection-induced humoral response in infants and young children and to evaluate potential differences with the adult response, we next tested the anti-spike IgG subclasses usage in a subset of samples representing early (30 – 140 days post infection) and late (250 – 375 days post infection) time points following infection (**Figure 4A**). The IgG responses can be further categorized into IgG1, IgG2, IgG3 and IgG4 antibody responses. Each of these subclasses have unique properties and effector functions that are driven by the Fc portion of the antibody molecule (22). Thus, we sought to characterize the IgG subclass usage of the SARS-CoV-2 antibody response. IgG1 was found to be the most dominant subclass in both infants/young children and adults, generating titers of 1 – 100 µg/mL in both infants/young children and adults. However, we observed notable differences in the anti-spike IgG subclass distribution. The second most prevalent IgG subclass in infants was IgG2, forming 1 – 10 µg/mL of the anti-Spike IgG response. In contrast, adults were significantly more likely to produce IgG3 (p=0.0294) (**Figure 4B**). These differences were most apparent at early time points, as the titers in adults dropped over time and titers in infants/young children maintained, confirming the higher durability of the antibody response in infants/young children.

**Figure 4:**
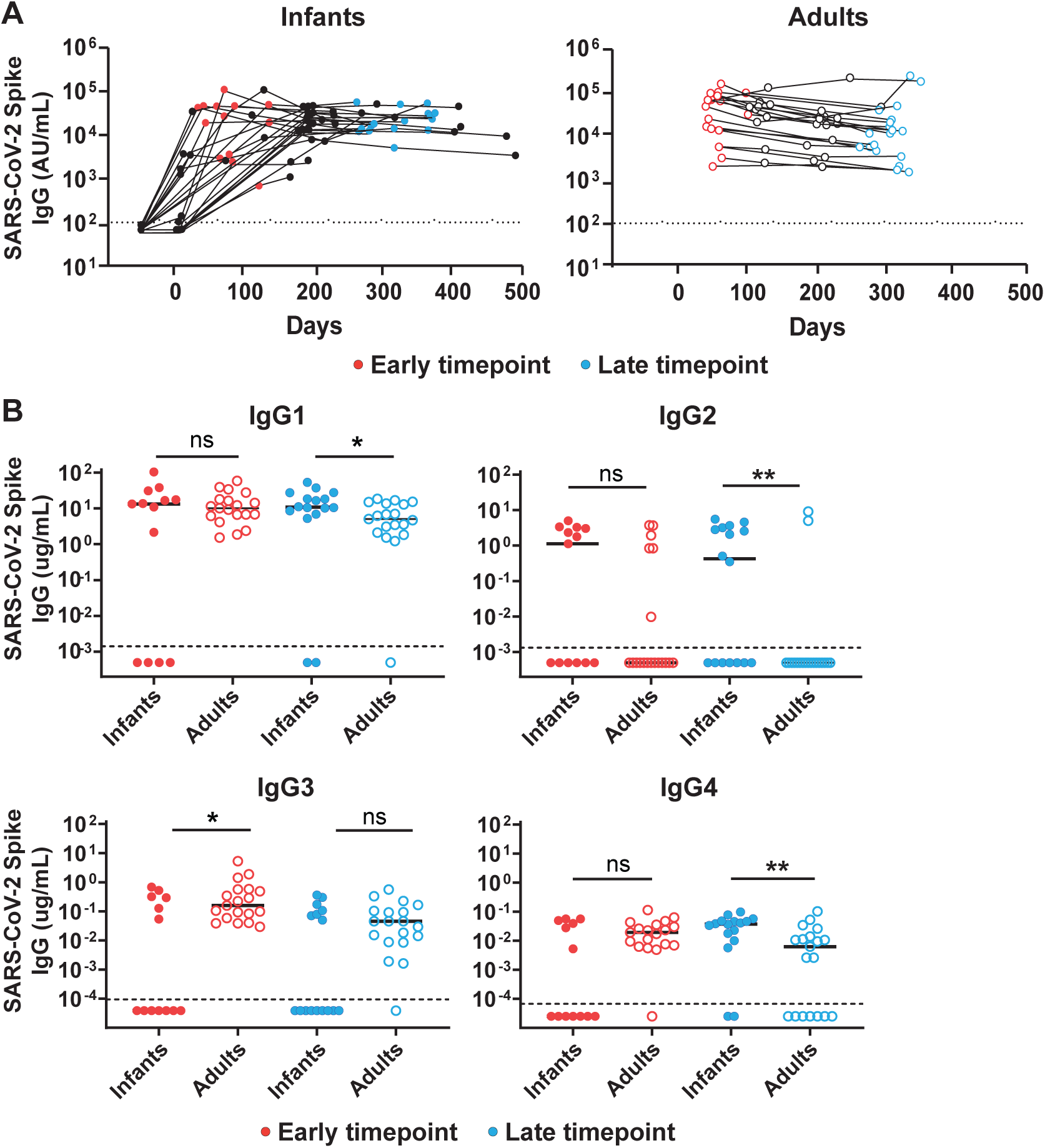
**Anti-spike IgG subclass distribution in infants/young children and adults.** The concentration of anti-spike IgG subclasses was measured at an early (30 – 140 days post infection, red) and late (250 – 375 days post infection, blue) time points. (A) Early and late time point samples from the infant and adult donors chosen to assess the antibody subclass distribution. (B) Concentration (ug/mL) of anti-spike IgG1, IgG2, IgG3 and IgG4 antibodies at early and late time points in infants and adults. Statistics were calculated using two-tailed Mann-Whitney test. ns – not significant, *p<0.05, **p<0.01.

### Infants/young children generate more durable neutralizing antibody response against WT SARS-CoV-2 as compared to adults

We next evaluated early and late time points in infants and adults following infection to assess the magnitude and durability of neutralizing antibody to WT (WA1\2020) SARS-CoV-2 strain (**Figure 5A**). The neutralizing antibodies were measured using a live virus focus reduction neutralization (FRNT) assay. Infants/young children and adults showed comparable neutralizing antibody titers against WT SARS-CoV-2 at the early post-infection time point, with median neutralizing antibody titers of 125 and 128 in infants/young children and adults, respectively. 73% of infants/young children and 68% of adults showed neutralizing antibody titer above the limit of detection (> 20). In contrast, at the late time point infants/young children showed a significantly higher neutralizing antibody response against WT SARS-CoV-2 as compared to the adults, with the median neutralizing antibody titers of 131 in infants/young children and 22 in adults (p = 0.0151). Furthermore, at the later time point 100% of infants/young children registered neutralizing antibody titer above the limit of detection as opposed to only 58% of adults.

**Figure 5:**
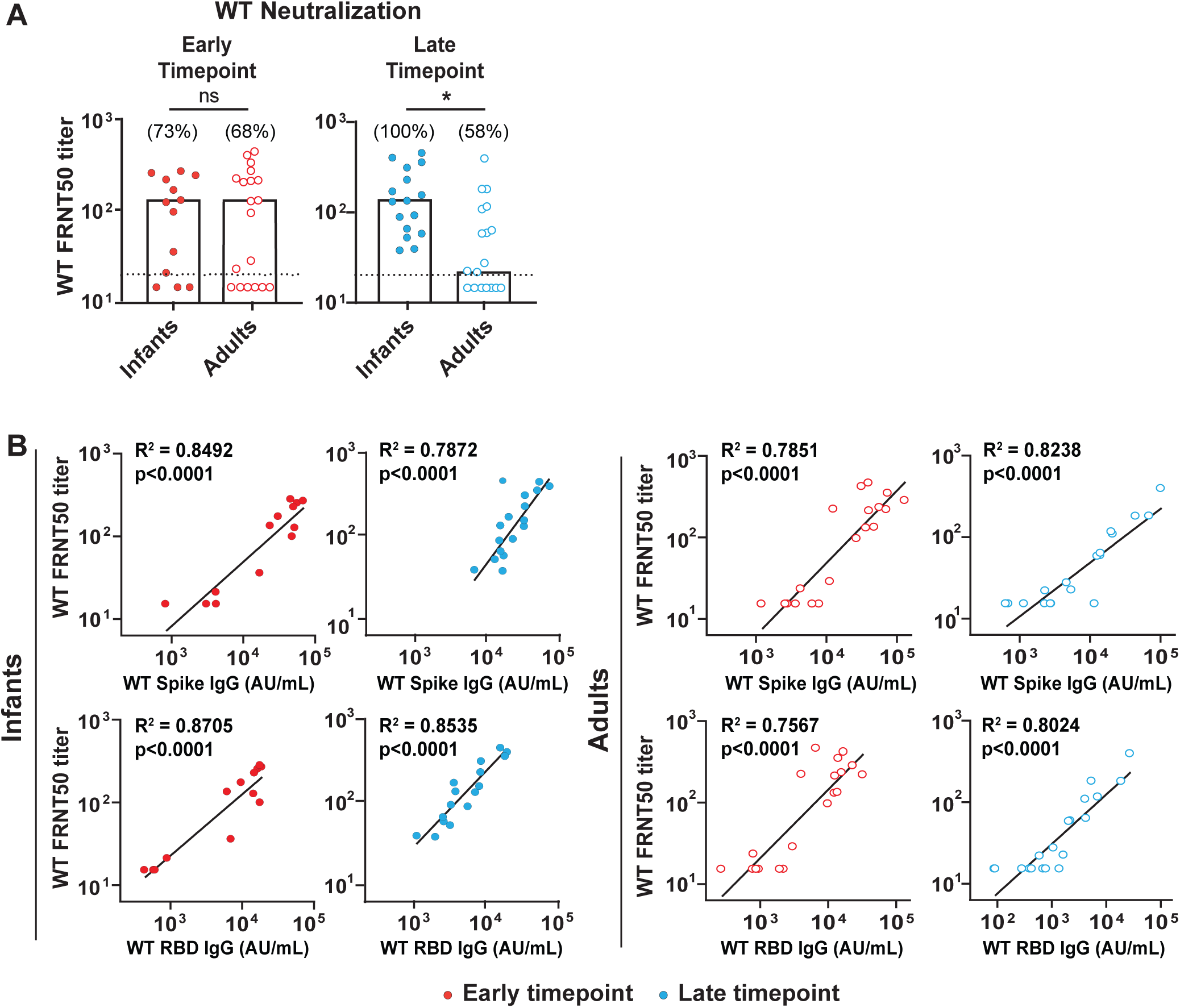
**Neutralizing antibody responses against WT SARS-CoV-2 spike in infants/young children and adults.** (A) Neutralizing antibody titers against WT SARS-CoV-2 at early and late time points in infants and adult donors. (B) SARS-CoV-2 spike and RBD reactive IgG (AU/mL) levels correlated with the neutralization titers at matched timepoints for infant and adult donors. The dotted line (A) represents the limit of detection at 1/20. The comparison between groups were performed using unpaired two-tailed t-test (A). The correlation analyses were performed using simple linear regression and coefficient of determination (R^2^) and significance were determined (B). ns – not significant, *p<0.05.

Next, we assessed the relationship between anti-spike and anti-RBD antibody and neutralizing antibody level in both infants and adults (**Figure 5B**). The spike and RBD IgG levels were found to correlate significantly to the neutralizing antibody titer, at both early and late time points, in infants/young children and adults (p < 0.0001) alike. The strong correlation of anti-spike and anti-RBD titers with neutralization, combined with the observation of increased durability of said titers, provides more evidence that infants/young children can maintain both binding and neutralizing antibody titers for a much longer period as compared to adults.

### The antibody breadth against VOCs is similar in infants/young children and adults

To address potential functional differences in the antibody repertoire, we analyzed the ability to bind and neutralize a panel of viral VOCs using early and late time point plasma samples from infants/young children and adults. These analyses included the beta, delta, and omicron (BA.1) variants. For the RBD IgG binding antibody response, infants/young children showed a 3-fold reduction and adults showed a 4-fold reduction in binding to beta variant. Infants/young children and adults showed a comparable 1 – 2-fold reduction in binding to delta variant. The reduction in binding was more pronounced in case of Omicron (BA.1) variant with infants/young children showing 17-fold reduction at early and 8-fold reduction at late time point, while adults showing 12-fold reduction at early and 9-fold reduction at late time point (**Figure 6A**). Similarly, for anti-spike IgG titers, infants/young children and adults showed a 2-fold reduction in binding to beta and 1 – 2-fold reduction in binding to delta variants. The reduction in binding was more evident in case of Omicron (BA.1) variant. The infants/young children showed a 10-fold reduction in binding at both early and late time points, while adults showed a 9-fold and 7-fold reduction in banding at early and late time points respectively (**Supplementary Figure 2**).

**Figure 6:**
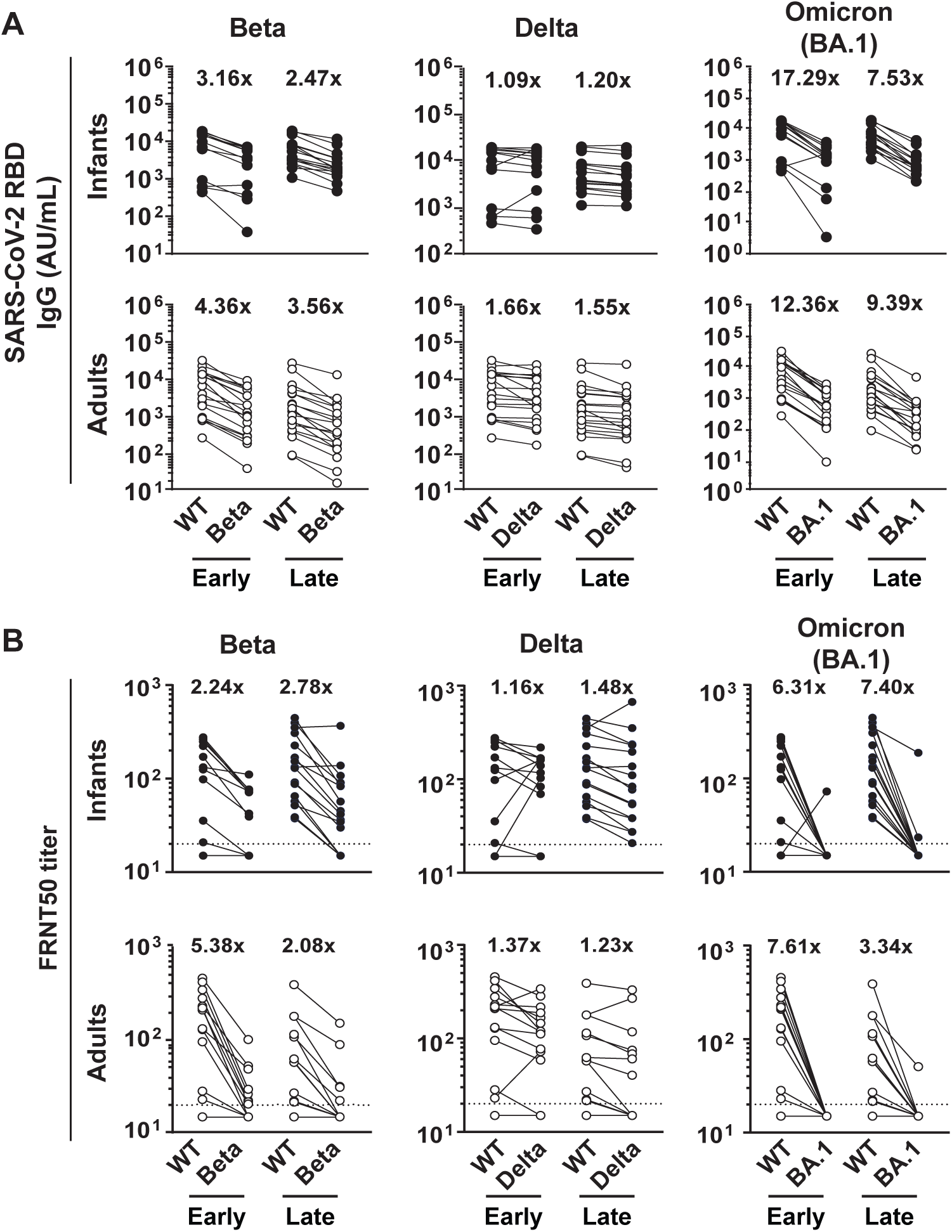
IgG binding and neutralizing antibody titers against WT, Beta, Delta and Omicron (BA.1) SARS-CoV-2 RBD in infant and adult donors. Infants exhibit similar breadth against VOCs as adults. (A) IgG binding to RBD and (B) neutralizing antibody titers (FRNT50) were analyzed against a panel of VOCs including WT, beta, delta and omicron (BA.1). The dotted line represents the limit of detection at 1/20 (B). Both infants’ and adults’ plasma samples were compared at early and late time points and fold change was calculated as variant binding (A) or neutralization (B) compared to WT.

Furthermore, both infants/young children and adults showed a significant reduction in neutralizing ability against beta, delta and Omicron (BA.1) variants. The infants/young children showed a 2 – 3-fold reduction in neutralization against beta and 1-fold reduction in neutralization against delta variant. Adults showed a 2 – 5-fold reduction in neutralization against beta and 1-fold reduction in neutralization against delta variant. The reduction in neutralization was even more evident in case of Omicron (BA.1) variant. Infants/young children showed 6-fold and 7-fold reduction, whereas adults showed 8-fold and 3-fold reduction in neutralization against Omicron (BA.1) at early and late time points respectively (**Figure 6B**). Taken together, these data suggest that the SARS-CoV-2 infected infants/young children and adults show a similar breadth of antibody response against VOCs, with a significant reduction in the binding and neutralizing ability against an Omicron (BA.1) variant.

## Discussion

As SARS-CoV-2 establishes itself as an endemic infection, exposure in early childhood may become routine. Therefore, it is critical to evaluate the magnitude and durability of immunity following early life infection, which may help to predict whether early immunity is sufficient to prevent new infections and severe disease later in life. Additionally, comparison of the immune response to infection in infants/young children to that of adults provides essential context for effective scheduling of primary and booster vaccinations across all age groups in view of continuously emerging viral variants. Our study of longitudinally sampled infants/young children is uniquely suited to evaluate these factors. These donors were followed with weekly nasal swabs and timely blood collections beginning soon after birth, which allowed us to identify both symptomatic and asymptomatic respiratory infections and evaluate the immune response both prior to SARS-CoV-2 infection and longitudinally after the first infection in early life. In addition, our cohort of longitudinally followed, non-severe adult COVID-19 patients enables a direct comparison of the infection-induced immune response in these age groups. To address the question of the durability of immune responses following infection, we evaluated the half-life of antibodies to SARS-CoV-2 antigens. The most striking finding of our study was that the infants/young children generated a much more durable binding and neutralizing antibody response as compared to adults following SARS-CoV-2 infection. In adults, the half-life of IgG binding antibodies was in the range of 178-204 days as estimated by the exponential decay model. This finding is in line with the range our group and others have reported in the past (5, 17, 18). It is well established that the loss of binding titers results in a coordinated loss of neutralization titer (**Figure 5A**) (16, 23–25), most likely due to the strong correlation between spike and RBD binding titers and neutralization (**Figure 5B**) (11, 26), indicating that fraction of anti-spike/RBD IgG is able to block SARS-CoV-2 entry into cells. The relatively rapid decline in binding and neutralization titers in adults is one factor that drives the recommended frequency of vaccine boosters. Therefore, it is a valuable insight that infants/young children showed a much more durable antibody response with half-lives of 700 – 800 days, nearly four times that of adults (**Figure 3**). Though the early and adult immune systems differ in a myriad of ways (27), infants/young children were able to generate robust neutralization titers. Indeed, infants/young children and adults produced neutralization titers comparable in magnitude early during the response to infection. In contrast to the rapidly declining neutralization titers in adults, the infants/young children did not exhibit significant reduction in neutralization titers at late time points, highlighting the longer durability of immunity following infection in this age group. These differences between infants/young children and adults could be due to different immunological milieus during the induction of the responses during acute infection, which could result in differences in the generation of long-lived plasma cells or be mediated by differences in the ability of the infant and adult bone marrow to support the survival of long-lived SARS-CoV-2 specific plasma cells. The mechanistic explanation for our findings requires further study. Notably, the striking differences in the durability of binding antibody to SARS-CoV-2 S antigens is not observed in regard to anti-nucleocapsid IgG. Infants/young children and adults exhibit a comparable rapid decline of anti-nucleocapsid IgG antibodies. This suggests that anti-spike, - RBD and -nucleocapsid humoral responses may be differentially regulated and implies that the increased durability in infants/young children is not necessarily universal for every antigen. It is possible that location, concentration, and/or availability of antigens differs during infection in adults compared to infants/young children, and that exploring these differences may lead to uncovering factors that could modulate durability of humoral immunity in adults. The spike protein of SARS-CoV-2 is a critical target of mRNA vaccines (28, 29), and the higher durability of spike-binding and neutralizing antibodies in infants/young children raise a possibility of similar increased durability of the immune response to vaccination, which may further impact booster scheduling in this age group. This cohort of donors, as well as peers who have not experienced infection, will continue to be followed, allowing us to build on these results and address important questions regarding durability of immune response to primary and booster series of vaccination in infection-experienced and naïve children. Overall, these findings provide important insight into the humoral immune response in very young children in a world in which childhood SARS-CoV-2 infection may become endemic.

Our study also observed variability in the IgG subclass usage in response to infection in infants/young children as compared to adults. As we have shown previously in adults (30), we observed a predominance IgG1 antibodies in both infants/young children and adults. Indeed, while IgG1 formed the most dominant subclass, adults and children differed in their secondary subtype, showing significant contribution of IgG2 in infants/young children and IgG3 in adults. IgG2 is known to be associated with neutralization and non-inflammatory response, and is known to reduce the detrimental effect of antibody-dependent enhancement (ADE), whereas IgG1, IgG3, and IgG4 are associated with both neutralization and ADE effects (31, 32). The higher contribution of IgG2 antibodies in infants/young children may contribute to less severe disease and fewer deaths in this age group as compared to adults. While subclass differences were subtle, these findings may provide important clues for how the immune response to infection is regulated in adults and infants/young children, and continued investigation may have broader implications for future vaccine development that could target a specific subclass of antibodies.

Another relevant question is whether pre-existing immunity against the commonly circulating human coronaviruses (HKU1 and OC43) provides cross-protection against SARS-CoV-2 infection? Although cross-reactivity and/or cross-protection between SARS-CoV-2 and HCoVs has been reported (33–35), we found no evidence of a major impact of pre-existing seasonal HCoVs titers on the magnitude or quality of SARS-CoV-2 responses in infants/young children. Since virtually all adults have experienced infection with common human coronaviruses and will therefore have pre-existing titers (5), the study of infants/young children provides a unique opportunity to assess responses to SARS-CoV-2 without pre-existing exposure. While this study could not measure protection, the pre-infection baseline titers of HCoVs’ spike protein did not correlate with the SARS-CoV-2 spike convalescent IgG antibody titers, suggesting pre-existing antibody titers did not result in an increased humoral response to SARS-CoV-2. It is also evident from low to non-existent IgG and IgA titers against the SARS-CoV-2 antigens observed in the pre-infection samples in the infants/young children, suggesting that the SARS-CoV-2 infection and immune response is independent of the HCoVs’ serostatus. This is in line with other studies done in older children and adults where the cross-protection by HCoVs has not been reported (36, 37). These important observations again highlight the need for vaccination against SARS-CoV-2 in infants/young children despite the pre-existing immunity against seasonal beta-coronaviruses. However, likely due to the high degree of spike protein conservation (21), we did find an increase in the IgG binding antibody titers against previous endemic coronaviruses, MERS-CoV and SARS-CoV-1 upon SARS-CoV-2 infection in infants/young children. This finding is consistent with other studies where SARS-CoV-1 titers were found to increase upon SARS-CoV-2 infection and mRNA vaccination in adults (5, 38). Taken together, these results may have implications for a broader strategy for vaccination targeting multiple human beta-coronaviruses.

Despite the differences in durability, our study demonstrated a very similar breadth of antibody repertoire in infants/young children and adults against VOCs such as beta, delta, and omicron (BA.1). We found lower binding and neutralizing antibody titers against beta and delta variants as compared to the WT in both infants/young children and adults, with differences being most apparent in the beta variant. As expected, the largest differences were observed for the omicron variant (BA.1). Prior SARS-CoV-2 infection in adults has shown to not confer cross-protection against the omicron variant (39, 40) in adults, an observation that impacted the approval of the bivalent booster. However, this has not been fully characterized in infants/young children. As observed in adults, we found a marked reduction in binding and neutralizing antibody titers against omicron (BA.1) variant in infants/young children. This underscores the potential of this VOC to evade the infection-induced immune response and cause re-infections in infants/young children even in light of their improved antibody durability. Booster vaccination increases responses to omicron in adults (41), and may rescue lower responses in infants/young children and may be necessary to protect from continuously emerging viral variants.

SARS-CoV-2 remains a global public health threat even after years of devastating disturbances and loss. To date, vaccination remains the best possible strategy in the fight against COVID-19. Thus, development of the booster vaccines that target evolving viral variants and optimum scheduling of vaccinations are both important factors in preventing new cases and progression to severe disease. A critical aspect for effective scheduling of vaccinations is understanding how long immunity persists following infection in various age groups. Our study demonstrated that infants/young children produce a significantly more durable antibody response to SARS-CoV-2 S antigens following infection as compared to adults, a response that correlates with extended neutralization, which may have implications for scheduling of vaccination and boosters. In addition, we found that natural infection confers a similar breadth of antibody repertoire in these disparate age groups, indicating a need for further study on the effect of boosters for prevention of continuously evolving variants.

### Limitations of the study

The major limitation of this study was the small cohort size which did not allow us to study the sex related effects to the SARS-CoV-2 infection induced immune response in infants/young children. Several reports have described notable differences in the immune response to COVID-19 between males and females (42–45). Thus, it is possible that this could have impacted our findings. However, while not fully powered to address this, we found no difference in the immune response to SARS-CoV-2 infection in infants/young children comparing males and females (data not shown). Future studies with larger number of infants/young children in the study cohort would be needed to better address this question. In addition, while our study compared an immune response to infection in infants/young children and adults, these donors were infected at different times during the pandemic most likely with corresponding main circulating viral variants. While it would have been interesting to compare the infants/young children and adults matched with respect to time of infection, we did not have the longitudinally followed matched donors to evaluate this. However, a similar breadth of antibody repertoire against VOCs in infants/young children and adults highlight the relevance of our findings.

## Methods

### Participant sample collection

The enrolled infants/young children at Cincinnati children’s hospital were part of a larger study of influenza infection and vaccination in early life. The study population included infants/young children who were followed since birth with weekly nasal swabs and twice weekly symptom surveys to diagnose the respiratory infections and describe clinical presentations, respectively. The blood and saliva samples were collected from these donors prior to and acutely after influenza vaccination or infection and SARS-CoV-2 vaccination or infection with longitudinally collected convalescent blood samples following these events. The Atlanta study population included adult volunteers over the age 18 who were diagnosed with COVID-19 by a commercially available SARS-CoV-2 PCR test. Informed consent was obtained from all participants prior to conduct of study procedures. The participants provided a medical history of co-morbidities, presentation of SARS-CoV-2 infection onset and disease course. The serial peripheral blood samples were collected starting approximately 30 days after SARS-CoV-2 infection and thereafter at 3, 6, and 9 months. The participants were excluded if they were immunocompromised, HIV positive, had active hepatitis B or C virus infection, used immunosuppressive drugs for 2 weeks or more in the preceding 3 months, received blood products or immunoglobulin 42 days prior to enrollment, received convalescent COVID-19 plasma, or were pregnant or breastfeeding.

### Viruses and cell lines

VeroE6-TMPRSS2 cells were cultured as described previously (46) and used to propagate all virus stocks. nCOV/USA_WA1\2020 (WA1), closely resembling the original Wuhan strain was propagated from an infectious SARS-CoV-2 clone as described previously (47). The B.1.351 variant isolate was kindly provided by Dr. Andy Pekosz (John Hopkins University, Baltimore, MD), was propagated once to generate a working stock (46). hCOV-19/USA/PHC658/2021 referred to as B.1.617.2 variant) was derived from nasal swab collected in May 2021 (48). EPI_ISL_7171744 (BA.1) variant was propagated once to generate the working stock (49). All variants were plaque purified on VeroE6-TMPRSS2 cells and propagated once in a 12-well plate of confluent VeroE6-TMPRSS2 cells followed by expansion of the working stock in T175 flasks on confluent VeroE6-TMPRSS2 cells. All viruses used in this study were deep sequenced and confirmed as previously described (48).

### Multiplex electrochemiluminescence (ECLIA) Immunoassay

The levels of IgG, and IgA antibodies were measured using the multiplex plates by Mesoscale Discovery (MSD), following manufacturer’s instructions. The V-PLEX Coronavirus Plate 1 (K15362U) were used to analyze the binding antibody responses against WT SARS-CoV-2 and other human coronaviruses, while V-PLEX SARS-CoV-2 panel 24 (K15575U) and panel 26 (K15593U) were used to assess the binding antibody responses to additional VOC-derived spike and RBD respectively. Briefly the plates were first blocked using the 5 % solution of Bovine Serum Albumin (BSA) in 1X PBS and incubated at room temperature with shaking at 700 RPM for at least 30 min. Following incubation, the plates were washed three times using 1X DPBS/0.05 % Tween. The serum samples, diluted 1:5000 (for IgG), and 1:1000 (for IgA, and IgM), were added and the plates were incubated at room temperature with shaking at 700 RPM for 2 h. Following a wash, sulfo-tag anti-human detection antibody (IgG, IgA, IgM, IgG1 Fc, IgG2, Fc, IgG3 Fc, or IgG4 Fc) were added, and the plates were incubated at room temperature with shaking at 700 RPM for 1 h. After the subsequent wash, 150 uL/well of MSD Gold Read Buffer B was added to the plates and the plates were read immediately on the MSD instrument to measure the light intensity. The levels of the antibodies are reported as the arbitrary units/ mL (AU/mL) or µg/mL based on the normalization of the standard curve.

### Focus reduction neutralization assay

The FRNT assays were performed as described previously (46, 48, 50). Briefly, the plasma samples were diluted 3-fold in eight serial dilution in DMEM in duplicates, with an initial dilution of 1:10 in a total volume of 60 µL, followed by incubation with equal volume of respective SARS-CoV-2 variant. The antibody-virus mixture was then added to VeroE6-TMPRSS2 cells. After incubation, the antibody-virus mixture was removed and pre-warmed 0.85 % methylcellulose overlay was added to each well. After removal of the overlay, the cells were fixed with 2 % paraformaldehyde in PBS. Following fixation, permeabilization buffer (0.1 % BSA, Saponin in PBS) was added to the permeabilized cells. The cells were incubated with an anti-SARS-CoV spike primary antibody directly conjugated to Alexafluor-647 (CR3022-AF647). The foci were visualized on an ELISpot reader (CTL).

### Statistical analysis

Mixed effects exponential and power law models implemented in Monolix (Monolix, Lixoft) were used to analyze the waning of the antibody response (day 30 to day 500 in infants/young children and day 30 to day 350 in adult donors). The exponential and power law models were formulated as ordinary differential equations, 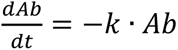 and 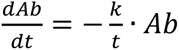, respectively, with antibody at day 30 lognormally distributed and decay rate *k* normally distributed and with lognormal multiplicative error. The data were analyzed using GraphPad Prism 9.5.0. The antibody neutralization titers were quantified by counting number of foci for each sample using the Viridot program. The neutralization titers were calculated as [1 – (average number of foci in wells incubated with patient serum) ÷ (average number of foci in wells incubated with control serum)]. The correlation analyses to determine coefficient of determination (R^2^), and significance were performed by log transforming the binding and neutralization titers followed by simple linear regression. The comparisons between groups were performed using unpaired two-tailed t-test.

### Study approval

The longitudinal IMPRINT influenza cohort study at Cincinnati Children’s Hospital Medical Center (Cincinnati, OH) and COVID-19 longitudinal cohort study at Emory University (Atlanta, Georgia) began after receiving the Institutional Review Board approvals (IRB 2019-0629, IRB00022371 respectively).

## Data Availability

All data presented in the manuscript will be available upon reasonable request to the corresponding author.

## Author Contribution

D.J. performed data acquisition, data curation and analysis, data visualization, and writing the manuscript. L.E.N and V.Z. contributed to data curation and analysis, data visualization, writing review and editing. A.M., K.M., and S.L contributed to investigation, data curation and analysis. A.R.B. and K.S. provided clinical support, contributed to patient recruitment and clinical data curation. G.M. and C.N. contributed to patient sample collection and paper review. R.A., E.J.A., M.A.S., M.S.S., and J.W. contributed to the conception and design of the study, writing and editing of the manuscript, and obtaining study funding. All authors have read and accepted the manuscript.

## Acknowledgements

We thank the participants and parents of infant donors for volunteering their time and efforts to participate in this study. We also thank Christina Rostad and Sanjeev Kumar for critically reviewing the manuscript. The graphical abstract was created using Biorender.com.

---

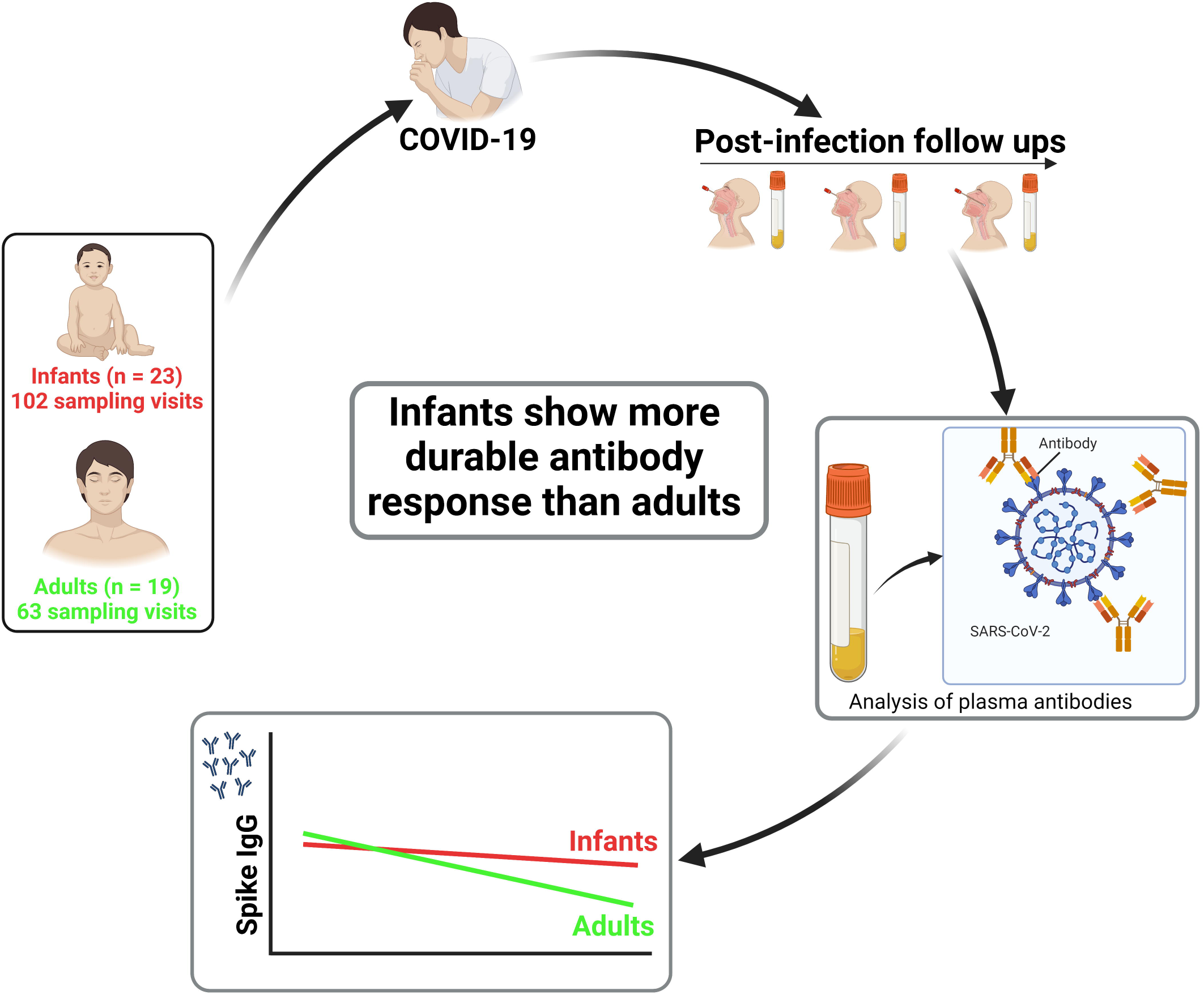

