## Supplemental Figures for "Infants and young children generate more durable antibody responses to SARS-CoV-2 infection than adults"

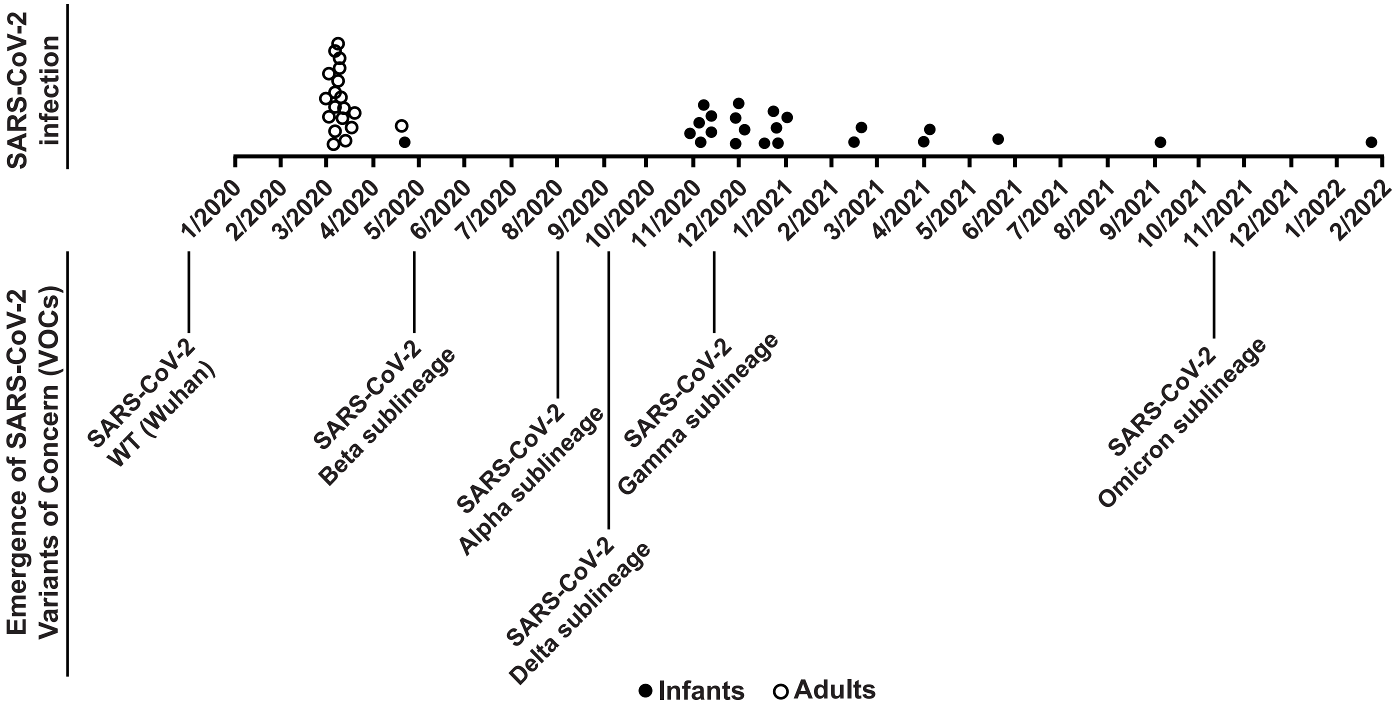


**Supplementary Figure 1: Infants and adults SARS-CoV-2 infection and currently circulating SARS-CoV-2 variants**

Time of infant and adult donors SARS-CoV-2 infection and corresponding circulating SARS-CoV-2 variants.

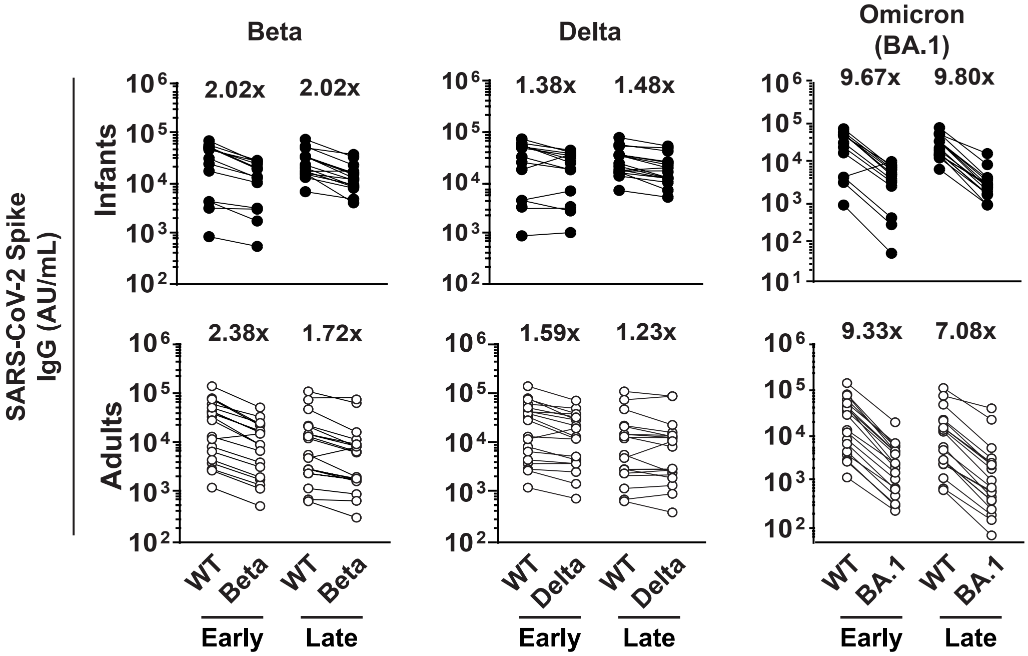


**Supplementary Figure 2: IgG binding antibody titers against WT, Beta, Delta and Omicron (BA.1) SARS-CoV-2 spike in infant and adult donors**

Binding antibody titers against spike were analyzed against a panel of VOCs including WT, beta, delta and omicron (BA.1). Both, infants' and adults' plasma samples were compared at early and late time points, and fold change was calculated.
